# Demographic and Viral-Genetic Analyses of COVID-19 Severity in Bahrain Identify Local Risk Factors and a Protective Effect of Polymerase Mutations

**DOI:** 10.1101/2022.08.13.22278740

**Authors:** Evan M. Koch, Justin Du, Michelle Dressner, Hashmeya Erahim Alwasti, Zahra Al Taif, Fatima Shehab, Afaf Merza Mohamed, Amjad Ghanem, Amani Alhajeri, Amna Alawadhi, Nabeel Almoamen, Khulood Ashoor, Sara Hasan, Alireza Haghighi, Shamil Sunyaev, Maha Farhat

## Abstract

A multitude of demographic, health, and genetic factors are associated with the risk of developing severe COVID-19 following infection by the SARS-CoV-2. There is a need to perform studies across human societies and to investigate the full spectrum of genetic variation of the virus. Using data from 869 COVID-19 patients in Bahrain between March 2020 and March 2021, we analyzed paired viral sequencing and non-genetic host data to understand host and viral determinants of severe COVID-19. We estimated the effects of demographic variables specific to the Bahrain population and found that the impact of health factors are largely consistent with other populations. To extend beyond the common variants of concern in the Spike protein analyzed by previous studies, we used a viral burden approach and detected a protective effect of low-frequency missense viral mutations in the RNA-dependent RNA polymerase (Pol) gene on disease severity. Our results contribute to the survey of severe COVID-19 in diverse populations and highlight the benefits of studying rare viral mutations.

## Introduction

As of April 2022, over 6.1 million deaths have been directly caused by severe coronavirus disease (COVID-19) after SARS-CoV2 infection (Ritchie et al. 2020). At the local epidemic level, the rate of severe cases is related to a confluence of factors including the circulating viral variants, and population characteristics ranging from demographics, to medical risk factors such as hypertension, diabetes, and genetics (Angioni et al. 2020, Ioannou et al. 2020, Williamson et al. 2020, Beaney et al. 2022, Kousathanas et al. 2022, Nyberg et al. 2022). A minority of studies have focused on severe COVID-19 in non-Western populations where mRNA vaccines have been less widely available (Booth et al. 2021). The systematic evaluation of demographic, medical, and viral risk factors for severe COVID-19 in different populations can improve our understanding of pathophysiology and inform policy around prevention moving forward. This is especially the case as it has recently become evident that existing vaccines do not adequately protect against mild infection and have limited ability to interrupt community transmission, necessitating a renewed focus on preventing severe COVID as the key outcome of public health interventions (Singanayagam et al. 2021).

Most analyses of viral genetics have focused on whether emerging variants, generally characterized by different combinations of Spike protein mutations, impact transmissibility and COVID-19 severity (Lin et al. 2021). For instance, the Alpha (B.1.1.7) variant has been associated with an increased mortality risk compared to previously circulating variants (Challen et al. 2021), while the Omicron (B.1.1.529) variant has been associated with a decreased mortality risk compared to the Delta (B.1.617.2) variant (Nyberg et al. 2022). Genome-wide association studies can be used to assess the severity-risk of every observed mutation in the SARS-CoV-2 genome (Hahn et al. 2021), but such an approach can lack power when the majority of mutations are at low frequencies and higher frequency mutations are grouped into emerging variants with strong temopral variation.

To move beyond existing work on common viral variants of concern in the Spike protein, we studied viral mutations that occur at lower frequencies and in other genes. Recent work has shown that many of the possible SARS-CoV-2 mutations have negative effects on viral fitness (Morales et al. 2021), and that these effects vary by gene (Lythgoe et al. 2021). However these mutations have a yet unknown impact on disease severity and COVID-19 outcomes.

Here, we prospectively analyzed data from 869 COVID-19 patients receiving inpatient or outpatient care in Bahrain between March 2020 and March 2021. We used paired viral sequencing and non-genetic host data to understand host and viral determinants of severe COVID-19 in Bahrain. Using multivariable regression and a viral burden approach, we detected a protective effect of rare missense viral mutations in the RNA-dependent RNA polymerase (Pol) gene, the target of the drug remdesivir, on disease severity. We find that rare mutations in Pol lead to a small but statistically significant increase in predictive ability for severe COVID-19, and that Pol is one of the most strongly constrained genes in the viral genome. Our results contribute to and support study of COVID-19 in diverse populations and highlight the benefits of studying rare viral mutations.

## Results

### Patient population

The median age of patients with COVID-19 was 37 years (IQR 27-51 years), and the majority of patients were male (59.5%). The Bahraini population is recognized to be majority male estimated at 64.7%, attributed to the sizable migrant worker population (The World Bank). The patient sample represented all four Bahraini Governates (Table 1). After Bahraini (58.4%), the next most common patient nationality was South Asian (18.2% spanning India, Bangladesh, Nepal, and Sri Lanka) (Table 1). The most commonly observed individual comorbidities were hypertension (16.6%), and diabetes (14.3%). A total of 30.2% of the patient sample were identified to have another comorbidity (Table 1).

**Table 1.** Baseline characteristics of COVID-19 positives.

| Table 1. Baseline characteristics of COVID-19 positives |  |  |
| --- | --- | --- |
| Characteristic | Patients (n=869) | Percentage |
| **Age group** |  |  |
| 0 - 20 years old | 94 | 10.8% |
| 21 - 40 years old | 410 | 47.1% |
| 41 - 60 years old | 248 | 28.5% |
| 61 - 80 years old | 99 | 11.4% |
| 81 - 99 years old | 18 | 2.1% |
| **Sex** |  |  |
| Female | 351 | 40.4% |
| **Nationality** |  |  |
| Bahrani | 509 | 58.6% |
| Missing | 70 | 8.0% |
| South Asia | 158 | 18.2% |
| Other | 132 | 15.2% |
| **Collection date** |  |  |
| Q1 2020 (March only) | 7 | 0.8% |
| Q2 2020 (May and June) | 100 | 11.5% |
| Q3 2020 | 71 | 8.2% |
| Q4 2020 | 191 | 22.0% |
| Q1 2021 | 497 | 57.1% |
| N/A | 4 | 0.4% |
| **Viral lineage** |  |  |
| 19A | 27 | 3.1% |
| 19B | 14 | 1.6% |
| 20A | 203 | 23.4% |
| 20B | 288 | 33.1% |
| 20I | 304 | 35.0% |
| Other | 33 | 3.8% |
| **Comorbidities** |  |  |
| Diabetes | 124 | 14.3% |
| Hypertension | 144 | 16.6% |
| Smoking | 88 | 10.1% |
| Other comorbidities | 101 | 11.6% |
| **Covid severity** |  |  |
| Severe | 106 | 12.2% |
| **Remdesivir** |  |  |
| Yes | 49 | 5.6% |
| **Region** |  |  |
| Capital | 273 | 31.4% |
| Muharraq | 133 | 15.3% |
| Missing | 147 | 16.9% |
| Northern | 219 | 25.2% |
| Southern | 97 | 11.1% |
| **Vaccination status** |  |  |
| Not vaccinated | 728 | 83.8% |

### Viral diversity

The earliest SARS-CoV-2 isolates (March-May 2020) belonged to clades 19A, 20A and 20B (Fig 1A). The clade composition shifted primarily to 20A and 20B in June 2021. The 20I (Alpha) clade appeared and rose in frequency in December 2021, coexisting with 20A and 20B for the duration of our sample. Numerous other clades were detected at low frequency and may represent the occasional importation of viral diversity from other countries. The first viral genetic principal component (PC) separated 20I (Alpha) from the remaining isolates (Fig 1B). The second partially separated 20A and 20B. Other PCs captured remaining genetic variation within clades 20A and 20B, and the seventh PC separated clades 19A and 19B from other clades (Fig. S1).

**Figure 1.**
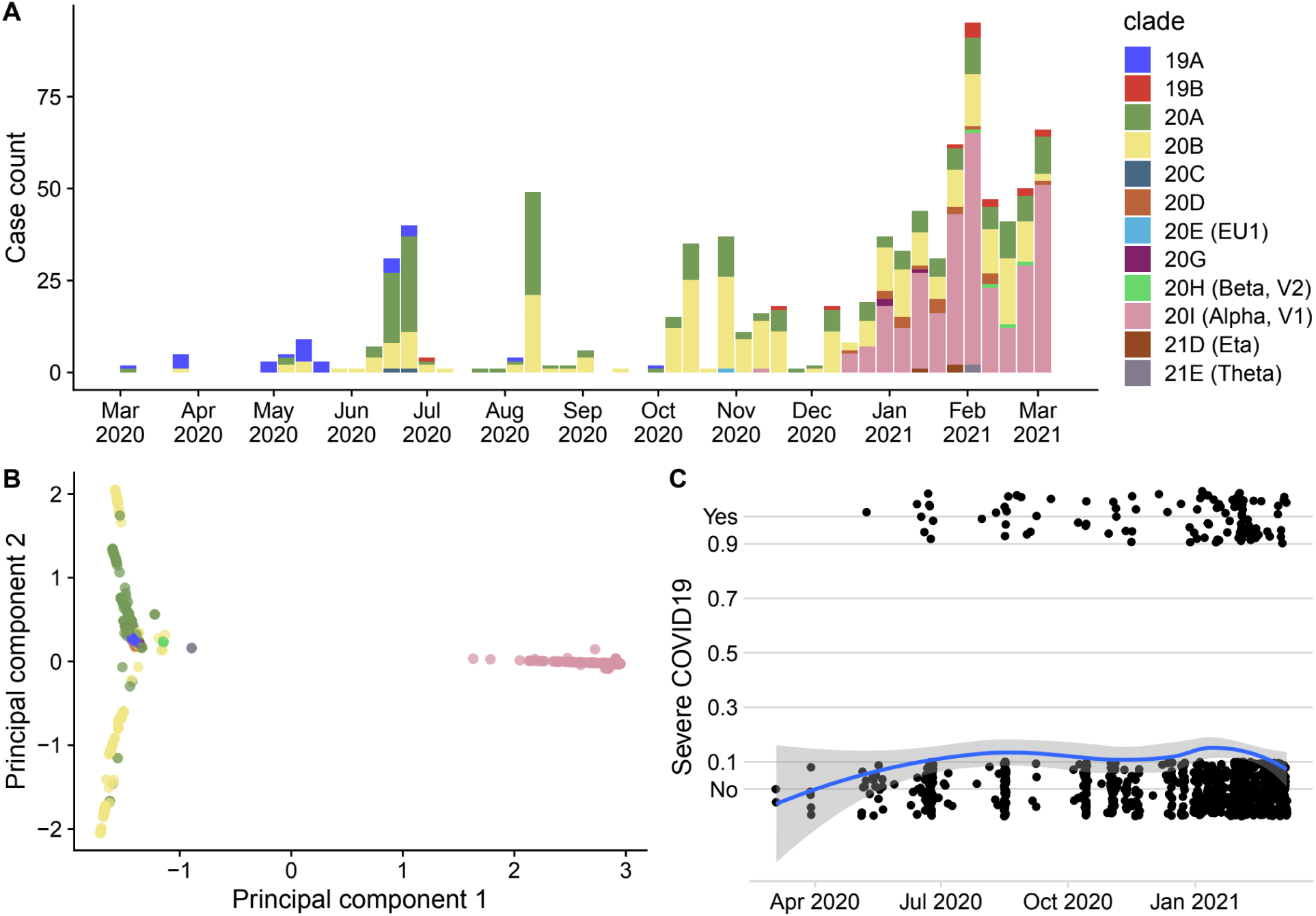
Summaries of the patient population (n=869) for which high-quality genomes and COVID-19 severity information were available. A) The distribution of cases and SARS-CoV-2 clades over the sampling period. B) The clustering of viral genomes on the first two principal components. The 20I (Alpha, V1) clade is separated from others along principal component 1. Principal component 2 captures variation within clades 20A and 20B, whose presence in the sample spans a longer time period. C) Individual cases and their COVID-19 severity are plotted as a function of sampling time. The blue line shows the LOESS-smoothed proportion of severe cases as a function of time.

### Host factors influence disease severity

In multivariable regression of host and socioeconomic factors (Table 2), vaccination with Sinopharm was found to decrease the odds of severe COVID-19 by 0.14 fold (95% CI=[0.05, 0.41]), and the odds of severe COVID-19 increased 2.11 fold (95% CI=[1.83, 2.44]) for each 10 year increment in age. Diabetes and other comorbidities were associated with, respectively, 2.36 (95% CI=[1.34, 4.15]), and 1.86 (95% CI=[1.01, 3.45]) higher odds of severe disease. Gender and sampling date were not significantly associated with severe disease, although the OR point estimate for female vs. male gender was <1 (Table 2). We did not observe an increased risk for severe disease with hypertension or smoking. Patients from the Muharraq governorate had decreased odds of severe disease (OR=0.36, 95% CI=[0.16, 0.79]) compared to the rest of the country, and individuals with a nationality other than Bahraini, or South Asian had increased odds (OR=2.52, 95% CI=[1.36, 4.64]).

**Table 2.**

| Value | n / median | Univariate OR | Univariate P-value | Adjusted OR | Adjusted p-value |
| --- | --- | --- | --- | --- | --- |
| Age (10 years) | 3.9 | 2.11 [1.83, 2.44] | 1.00E-23 | 1.97 [1.64, 2.36] | 5E-13 |
| Gender (Female) | 351 | 0.84 [0.52, 1.36] | 4.80E-01 | 0.70 [0.41, 1.18] | 2E-01 |
| Time (months) | 01/17/2021 | 1.05 [0.96, 1.14] | 2.80E-01 | 1.04 [0.93, 1.18] | 4E-01 |
| Vaccination status | 142 | 0.14 [0.05, 0.41] | 3.30E-04 | 0.14 [0.05, 0.42] | 5E-04 |
| Other Comorbid | 101 | 1.86 [1.01, 3.45] | 4.60E-02 | 1.81 [0.92, 3.57] | 9E-02 |
| Diabetes | 124 | 2.36 [1.34, 4.15] | 2.96E-03 | 2.44 [1.27, 4.70] | 7E-03 |
| Hypertension | 144 | 1.48 [0.84, 2.59] | 1.70E-01 | 1.17 [0.60, 2.25] | 6E-01 |
| Smoking | 89 | 0.84 [0.32, 2.10] | 6.90E-01 | 0.70 [0.26, 1.90] | 5E-01 |
| **Region** |  |  |  |  |  |
| Capital | 273 | - | - | - | - |
| Muharraaq | 133 | 0.36 [0.16, 0.79] | 1.12E-02 | 0.35 [0.15, 0.80] | 1E-02 |
| Northern | 219 | 0.86 [0.46, 1.59] | 6.20E-01 | 1.06 [0.55, 2.05] | 9E-01 |
| Southern | 97 | 0.83 [0.38, 1.83] | 6.50E-01 | 0.73 [0.31, 1.68] | 5E-01 |
| Missing | 147 | 0.86 [0.36, 2.07] | 7.40E-01 | 0.90 [0.32, 2.53] | 8E-01 |
| **Nationality** |  |  |  |  |  |
| Bahraini | 510 | - | - | - | - |
| South Asia | 158 | 0.68 [0.32, 1.43] | 3.10E-01 | 0.76 [0.35, 1.68] | 5E-01 |
| Other | 132 | 2.52 [1.36, 4.64] | 3.15E-03 | 3.52 [1.79, 6.93] | 3E-04 |
| Missing | 70 | 1.20 [0.44, 3.31] | 7.20E-01 | 1.55 [0.44, 5.42] | 5E-01 |

### Vaccination impacts the distribution of viral clades

We examined the relationship between vaccination and clade in our patient panel. Vaccination with Sinopharm started in Bahrain in November 2020. The COVID-19 clade composition in January 2021 has a relatively equal proportion of 20I (Alpha, V1) in previously vaccinated and unvaccinated COVID-19 patients. However, in February and March 2021, the proportion of 20I (Alpha, V1) isolates increased in vaccinated individuals relative to the unvaccinated to reach a ratio of approximately 2:1 (Fig 3). We found a significant association between 20I (Alpha, V1) and breakthrough COVID-19 among individuals vaccinated with Sinopharm compared with unvaccinated control COVID-19 patients matched by diagnosis date (OR=8.00, 95%CI=[4.06, 16.52], p=2E-11).

**Figure 2.**
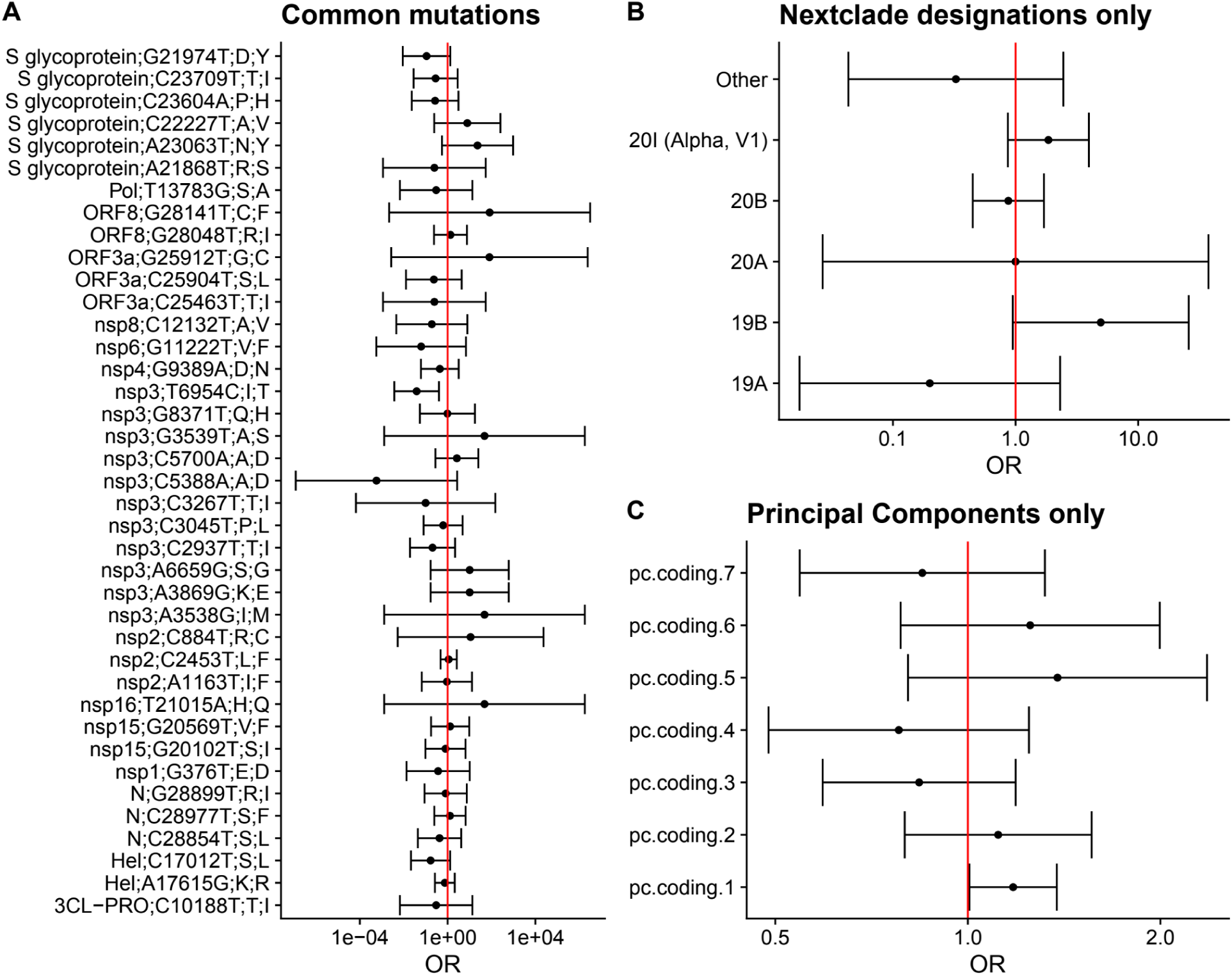
Results of analyses estimating the effects of common viral mutations, viral clades, and viral principal components on the probability of severe COVID-19 in patients. Effect sizes are given as odds ratios and 95% confidence intervals are displayed. A) The effects of the top 50 highest frequency mutations in the sample were estimated using logistic regression. 11 of these variants were collinear with other variables and were omitted. The remaining variants are labeled by the protein, the position in the reference genome, the reference/mutant nucleotide, and the reference/ mutant amino acid. Nextclade designations and principal components were both included in the common mutation analysis. B) The effects of Nextclade clade designations on COVID-19 severity. C) The effects of the top seven principal components on COVID-19 severity. Odds ratios are per-unit in principal component space.

**Figure 3.**
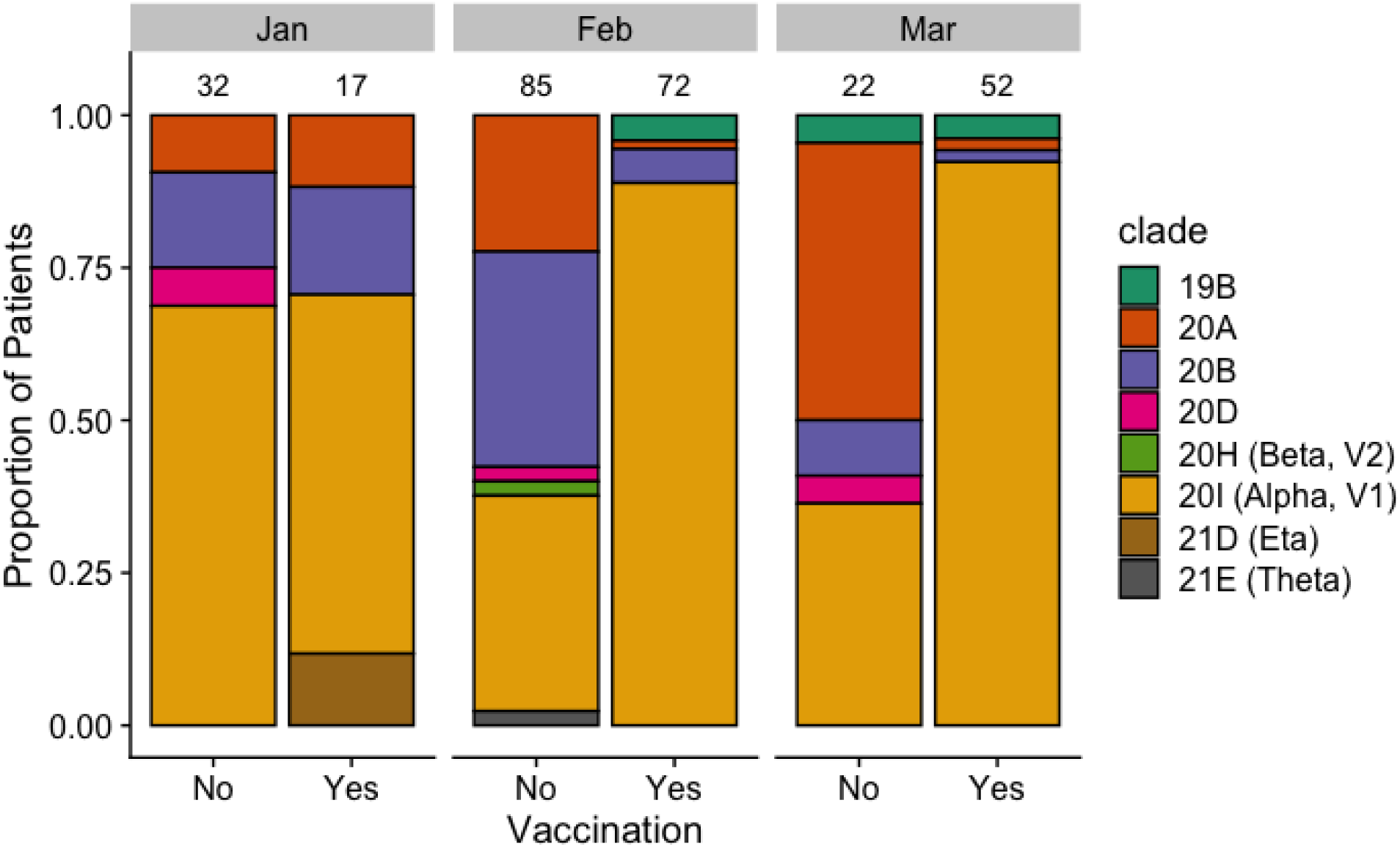
The proportion of different clades among vaccinated and non-vaccinated individuals in the final three months of the sampling period. Only clades actually observed during this period are shown. Numbers above bars indicate the patient counts in the vaccinated and unvaccinated categories.

### Viral factors influence disease severity

To study the effect of viral genetics on severity, we assessed the effects of common mutations controlling for population structure using the principal components as fixed effects. No common mutations were significantly associated with COVID-19 severity at the Bonferroni-corrected threshold of 0.001 (Fig 2A). One potential reason for this is that relevant patterns of common genetic variation were captured in the clade designations and principal components (Figs 2B and 2C). We next tested individually the effect of clades or PCs (analyzed separately due to high collinearity). Individually both clade 20I (Alpha, V1), and PC1 were associated with increased risk of severe COVID-19 with OR 1.85 (95% CI=[0.87, 3.96]), 1.18 (95% CI=[1.0, 1.38]) respectively.

We investigated low-frequency mutations and their relationship with COVID-19 severity. We first aggregated rare missense and rare synonymous mutations across all genes and found that neither score had a strong or significant effect on severity (Fig 4A). To account for any overall confounding signal of rare variation, we included synonymous burden scores as a covariate in missense analyses (Methods).

**Figure 4.**
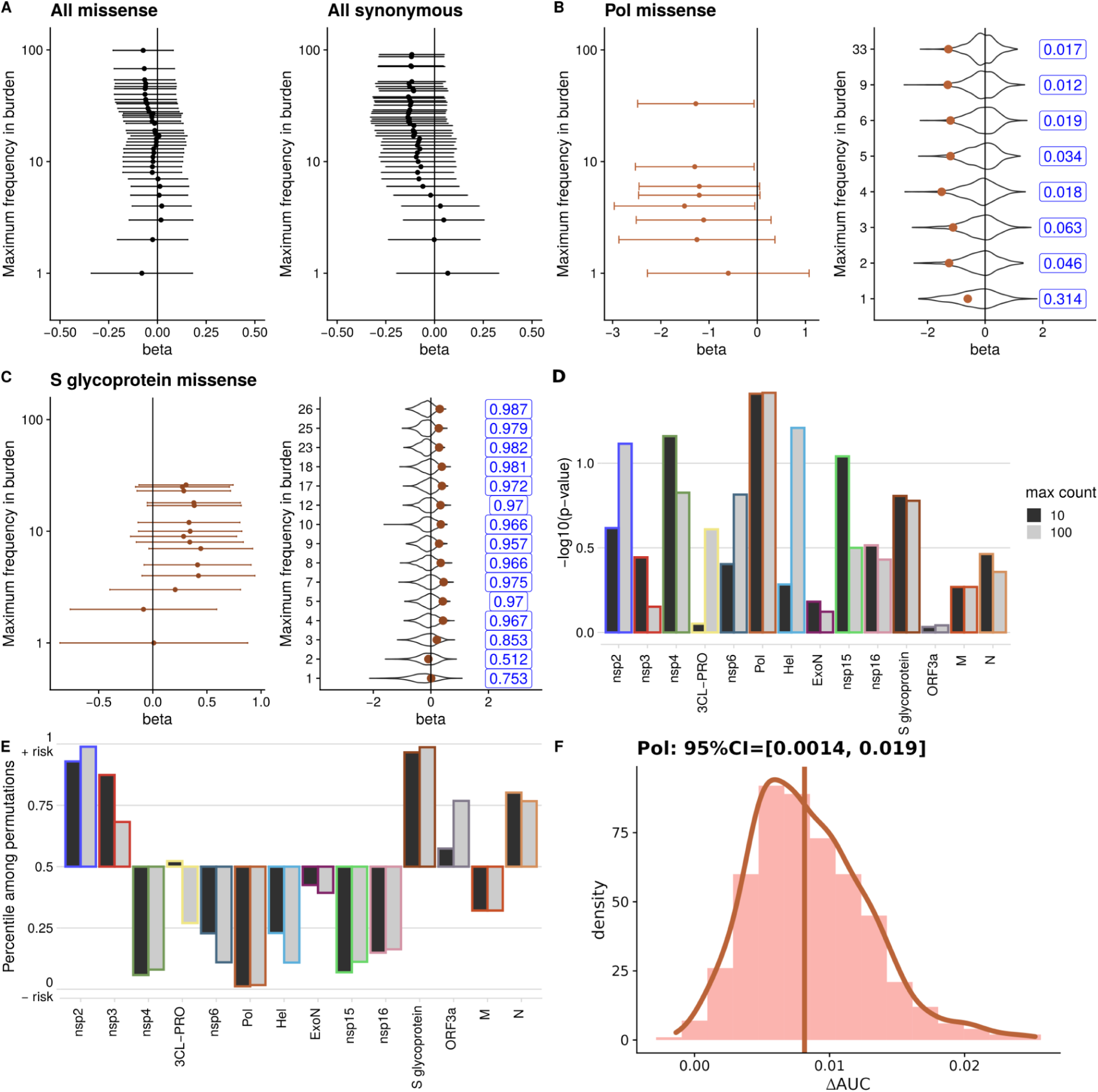
Results of burden tests for the association between low-frequency SARS-CoV-2 missense mutations on COVID-19 severity. Association results are presented as logistic regression coefficients (beta) per-mutation in the burden score. 95% confidence intervals are also shown. Violin plots show the distribution of coefficients from regressions calculated on randomized burden scores holding variant frequencies constant. Numbers to the right of violin plots give the percentile of the observed coefficient in the randomized distribution. A) Results for burden scores calculated on all missense and all synonymous mutations below each maximum value. B-C) Results for missense mutations in the Pol and S glycoprotein proteins. D) P-values of burden scores across all analyzed proteins at maximum counts of 10 and 100. E) Percentiles of estimated coefficients at maximum counts 10 and 100 in randomized distributions for all analyzed proteins. F) Changes in AUC resulting from adding Pol max-10 burden score to predictions of COVID-19 severity using logistic regression. Vertical line shows the 𝚫AUC=0.008 95%CI=[0.001, 0.017]), no prediction improvement adding AUC calculated by applying LOOCV to the full patient data. Distributions show 𝚫AUC=0.008 95%CI=[0.001, 0.017]), no prediction improvement adding AUC values calculated by applying LOOCV to bootstrap samples from the patient data. 95% confidence intervals are calculated from bootstrap samples.

**Figure 5.**
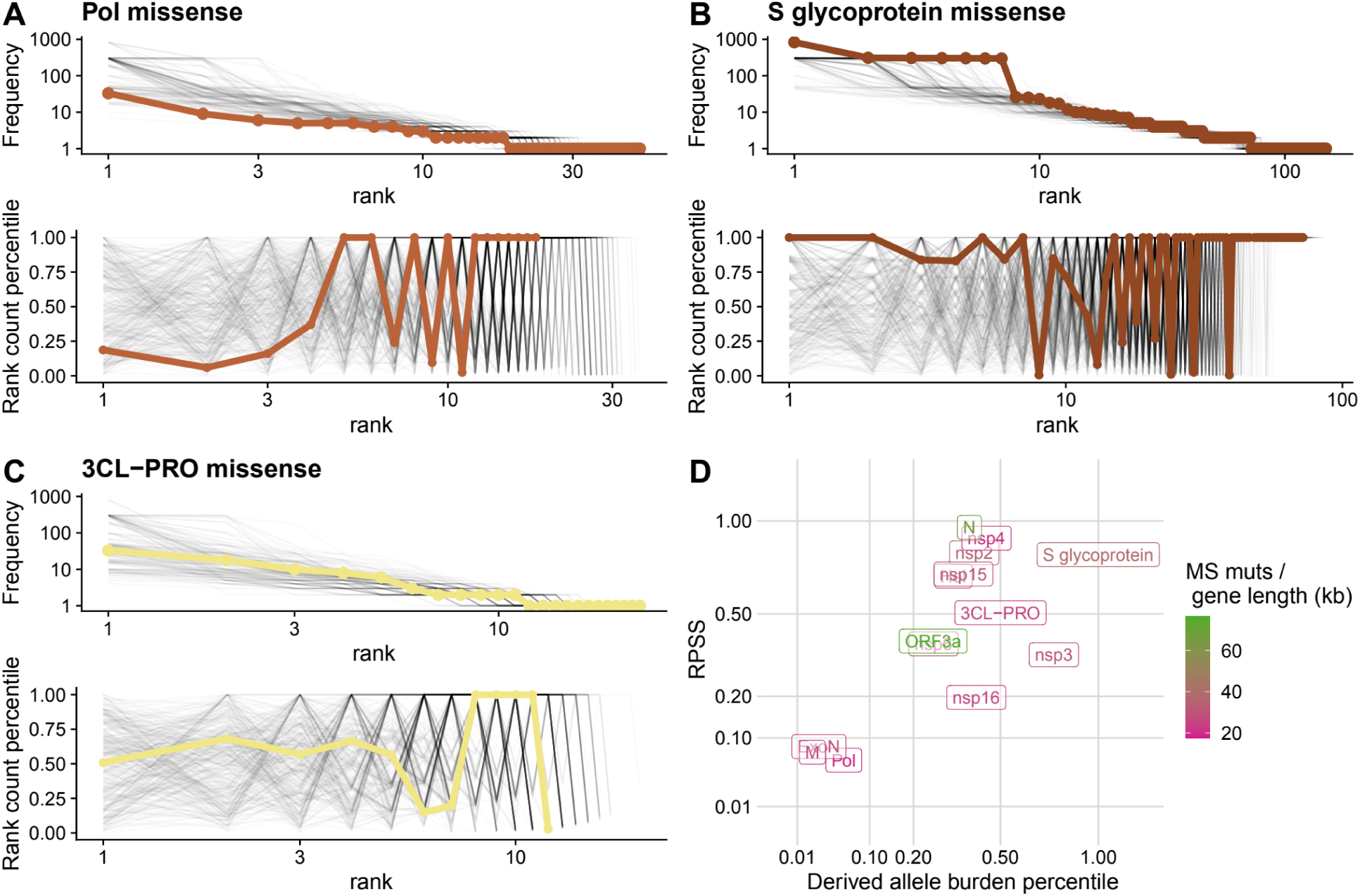
A-C) Distributions of observed SARS-CoV-2 mutation frequencies in genes of interest. Top plots show the frequencies of each mutation observed in a given gene, ordered by their rank. Gray lines represent random draws of missense mutations from the SARS-CoV-2 genome with the same number of mutations as observed in that gene. Bottom plots show the percentile of each mutation in a gene with respect to the distribution of variants genome-wide with frequency lower or equal to the mutation with the next lowest rank. Gray lines show these percentiles for the simulated set of mutations. Lines truncate when the frequency reaches one. The Rank Percentile Sum Score (RPSS) is the percentile of the area under the bold line against randomizations. D) The RPSS is plotted against the derived allele burden percentile of each gene. The derived allele burden is the average number of missense mutations in a gene, and the percentile is compared to randomizations with the same number of mutations.

We decided to test the hypothesis that rare variants in key genes collectively may result in reduced (or altered) COVID-19 severity. We decided to focus on two obvious primary candidates: S glycoprotein (Spike) because of its role in stimulating neutralizing immunity and RNA-dependent RNA polymerase (Pol) because of its role in remdesivir resistance. There is no signal in Spike, but there is a biologically plausible nominally significant signal in Pol. We found a significant protective effect of rare mutations in the Pol gene (max-freq:10, per-mutation OR= 0.27 [95% CI 0.08, 0.94, p=0.039]; max-freq:100, per-mutation OR= 0.28 [95%CI 0.084, 0.93, p=0.038]), and no significant effect of the rare mutation burden in the Spike gene (max-freq:10, per-mutation OR=1.41 [95%CI 0.88, 2.27, p=0.16]; max-freq:100, per-mutation OR= 1.36 [95%CI 0.88, 2.10, p=0.17]) (Figs 4B and 4C). The association with decreased severity in Pol was robust to using either clades or PCs as covariates (Fig. S2).

Motivated by this result, we then tested burden scores in the remaining 12 genes in the SARS-CoV-2 genome containing ≥200 amino acids. None were significantly associated with COVID-19 severity at the nominal threshold of 0.05 (Fig 4D, Fig S2). Thus, Pol is the only gene that harbors rare alleles apparently reducing disease severity. We note, however, that this result would not have been genome-wide significant using a Bonferonni correction and reflects our original focus on a biological hypothesis.

We also assessed the association of each gene with disease severity using a permutation score in which we determined the percentile of a gene-specific burden score relative to the distribution of burden scores for randomly chosen mutations at matched frequencies. This was done to ensure that it was not somehow the particular combination of variant frequencies driving severity associations. Pol burden scores were at the 1.2 and 1.7 percentiles of simulations for maximum frequencies of 10 and 100 respectively (Fig 4B). Spike burden scores were at the 96.6 and 98.7 percentiles compared to simulations (Fig 4C). We note that as the background distribution of mutations was biased towards risk-decreasing effects, and Spike mutations appear relatively risk-increasing in that context. Missense burden scores in all other genes had percentiles less extreme than Pol and Spike (Fig 4E).

Another way to summarize the association between genetic variables and COVID-19 severity is by their ability to improve prediction accuracy relative to a model with only host characteristics. We quantified the improvement in prediction accuracy by the change in AUC in a leave-one-out cross-validation (Methods). We found a modest improvement in severity prediction adding the Pol burden score (𝚫AUC=0.008 95%CI=[0.001, 0.017]), no prediction improvement adding AUC=0.008 95%CI=[0.001, 0.017]), no prediction improvement adding scores for Spike or principal component 1 to the host characteristics model (𝚫AUC=0.008 95%CI=[0.001, 0.017]), no prediction improvement adding AUC=0.0009, 95%CI=[-0.003, 0.01]) (Fig. S3).

### Signatures of selection on the SARS-CoV-2 genome

SARS-CoV2 genes varied widely in their diversity, ranging from 76.4 missense mutations per kb for the ORF3a protein to 17.5/kb for Pol (Fig 5D). To compare genes we computed a rank percentile sum statistic (RPSS, Methods) which evaluates whether the ranked mutations observed in a gene tend to decrease in frequency faster or slower than the rest of the genome. This statistic was designed to be less sensitive to the presence of outlier high-frequency and potentially positively selected mutations. In addition to having low diversity across the sample, the Pol gene (Fig 5A) had the lowest RPSS score (0.061) among genes longer than 200 amino acids, and only the Exonuclease (ExoN) and M genes were at lower percentiles in the simulated distribution of derived allele burdens. The Spike gene (Fig 5B) had the highest derived allele burden percentile (0.999) and the fourth highest RPSS score (0.799). Other genes such as the 3-C like protease (3CL-PRO) fell towards the middle of the distribution of mutation frequencies, and might be considered as under typical selection pressure for the SARS-CoV-2 genome (Fig 5C). The derived allele burden and RPSS measures of selection were correlated among genes (Fig 5D).

## Discussion

The development of severe COVID-19 is determined by patient demographics and medical risk factors in a complex interplay with viral and host genetics. In this study we sough to disentangle this interplay in the Bahraini population by using a large dataset enriched for severe COVID-19 with available viral sequences and metadata on relevant non-genetic host risk factors. We reproduced the expected impact of age and comorbidities on COVID-19 severity. However, we did not find a significant effect of gender on COVID-19 severity, whereas other studies have found that the fatality rates are higher for men than in women (Beaney et al. 2022), risk differences by gender have been shown to vary considerably with geography and time, suggesting that this interaction is mediated by social rather than biological factors (Danielsen et al. 2022) and highlighting the benefits of studying epidemics in different populations.

That we found an increased risk of severe COVID-19 among certain non-citizen residents of Bahrain (‘Other’ category, Table 2), and within one of four governantes (Muharraq) indicates some degree of demographic heterogeneity of risk within the Bahraini population. Interestingly, the fact that South Asian patients had no detectable increased risk of severe disease, and that many South Asians in Bahrain are migrant workers, may mean that correlates of migrant worker status were not important determinants of COVID-19 outcomes. This is possibly due to a younger age distribution and the general availability of treatment. Overall, because our analysis was restricted to the relatively small number of patients for which viral genetic data were available, we were underpowered to detect associations with demographic and medical risk factors with more subtle effects.

While it was important to assess the impact of non-viral factors, the major goal of this study was to investigate the association between viral genetic variation and the risk of severe COVID-19. Global genetic surveillance of the SARS-CoV-2 pandemic has demonstrated the emergence of competing successive strains that regularly displace one another. During our sampling period this included the near replacement of the original genotype with clades carrying the D614G mutation in the Spike gene (Plante et al. 2021), and then the rise in frequency of the Alpha strain (Walker et al. 2021). While these large-scale genetic changes can causally impact disease severity, they can also be correlated with different segments of the population becoming infected or with changing medical treatment. It takes careful study design to confidently assess the impacts of large-scale shifts in viral genetics on host outcomes.

A previous genome-wide association study (GWAS) of SARS-CoV-2 used a low-dimensional representation of the genetic similarity matrix to control for the effects of large-scale genetic shifts in the virus (Hahn et al. 2021). We took a similar approach and used clade designations (Hadfield et al. 2020; Aksamentov et al. 2021) and principal components of the genotype matrix. While we did detect a borderline-significant association between 20I (Alpha, V1) and increased risk of severe COVID-19, there was not a strong overall effect of either clades or principal components (Fig 2B and 2C). One possible reason for this is that variation in clade frequencies is strongly time-dependent and confounded with other time dependent changes in prevention and treatment. We attempted to control for temporal changes by including a time lapse variable, as well as using a sampling strategy that kept the proportion of severe cases relatively constant (Fig 1C). At the same time reducing the temporal component coould attenuate our ability to detect the effects of large-scale shifts in viral genetics.

In contrast to the GWAS approach, which tests every single observed mutation in the SARS-CoV-2 genome (Hahn et al. 2021), we used a burden approach to test individual genes. We initially tested the RNA-dependent RNA polymerase (Pol) and S-glycoprotein (Spike) genes, as these were large genes with the greatest prior evidence for affecting COVID-19 severity. Pol is the target of the drugs remdesivir (Kokic et al. 2021) and molnupiravir (Kabinger et al. 2021), contains many domains conserved across RNA viruses (Xu et al. 2003), and mutations in Pol have been shown *in vitro* to affect viral fitness (Szemiel et al. 2021) as well as the response to remdesivir treamtent (Gandhi et al. 2022). The Spike protein contains the receptor binding domain necessary for binding to ACE2 and subsequent entry into human cells. Mutations in Spike are capable of altering this binding affinity (Starr et al. 2020). By first testing these two genes and then performing an exploratory analysis on the remaining SARS-CoV-2 genes over 200 amino acids we drastically reduce the multiple testing problem compared to analyzing the whole genome.

Our main finding is that rare mutations in the Pol gene decrease the risk of severe COVID-19 (Fig 4A). Although the confidence intervals of the estimated effect are wide, ranging from 5% to 90% decrease in risk per rare mutation, the signal was robust to different approaches to controlling for population structure (Fig. S2), and was below the 2nd percentile of permuted mutations with the same frequency against a genomic background biased towards protective effects (Fig 4B). The inclusion of a Pol burden score significantly increased the predicted value of a logistic model. The magnitude of this effect was around a 1% increase in AUC. This modest improvement is expected given that the baseline probability of severe covid is 12% in our sample, and 10% of patients had at least one mutation with a Pol mutation of count 10 or lower.

Additionally, genetic variation in the Pol gene had one of the strongest signals of negative selection in the SARS-CoV-2 genome, a finding that agreed with the analysis of intra-host genetic variation in a previous study (Lythgoe et al. 2021). Phylogenetic analyses of sarbecoronaviruses as well as global variation within SARS-CoV-2 have indicated that much of the genome is under predominantly negative selection (Jungreis et al. 2021, Morales et al. 2021, Ghafari et al. 2022). We looked for differences in the strength of negative selection between genes by comparing the frequencies of missense mutations in our sample. Despite selective pressure to keep them from spreading, mutations with negative impacts of viral fitness can drift to detectable frequencies through stochastic superspreader and founder events (James et al. 2007). Deleterious mutations can also hitchhike to higher frequencies when they occur on relatively more fit genetic backgrounds. The overall expectation is that more frequent mutations have less negative effects on viral fitness compared to less frequent ones. By extension, genes with mutations at lower frequencies are more likely to be prone to mutations with negative viral fitness consequences.

In contrast to Pol, the burden of rare mutations in the Spike gene was not significantly associated with changes in the risk of severe COVID-19 although it was found to be in the upper percentiles of possible mutation permutations in increasing the risk of severe COVID-19 (Figs 4C, 4D, and 4E). Clade-defining Spike mutations have been repeatedly characterized to be under positive selection and associated with immune-escape and/or transmissibility but not consistently with changes in disease severity (Rochman et al. 2021). Mutations that either increase or decrease ACE2 binding affinity in Spike have been identified experimentally (Starr et al. 2020), but whether these increase the risk of severe COVID-19 in the hosts where they appear remains unknown.

An unanswered question is whether negatively selected mutations like the mutations observed in Pol, will increase, decrease, or have no effect on the risk of severe disease. Viral fitness is determined by a combination of the ability to survive and replicate with hosts, and the ability to transmit to new hosts. The first, within-host component of fitness may be related to disease severity. Our results support the idea that mutations in Pol, as the central component of viral replication, are likely to affect this fitness component. Future studies with large sample sizes, analyzing currently circulating strains of SARS-CoV-2, will be necessary to conclusively demonstrate the importance of rare mutations for COVID-19 outcomes.

## Methods

### Study population and data collection

We prospectively collected, through the chart abstraction from the Bahrain Ministry of Health (BMOH) medical records, demographic and health information for 1,151 patients in Bahrain with PCR-confirmed SARS-CoV-2 infection between March 4th, 2020 and March 8th 2021. These SARS-CoV-2 infected patients were chosen for sequencing based on (1) sampling to evenly represent asymptomatic or mildly symptomatic SARS-CoV2 patients across the different Bahraini geographic districts, or (2) the development of severe COVID as defined by an O2 saturation <90% or PaO2 <60%, and a physician determined need for ICU hospitalization, and the presence of respiratory disease including ARDS, or viral pneumonia or the need for mechanical ventilation or death due to COVID-19. Host level characteristics including comorbidities were abstracted from the patient electronic health records and physician questionnaires. These variables were then cleaned by correcting misspellings, categorizing fields as needed, and ensuring all null and missing values were entered as detailed in the code accompanying this paper.

### SARS-CoV-2 Sequencing

Viral RNA underwent cDNA synthesis using Invitrogen SuperScript IV VILO (SSIV VILO) Master Mix. Enrichment of SARS-CoV-2 genome was done via PCR using NEB Q5 High-Fidelity DNA Polymerase and ARTIC SARS-CoV-2 primer pools. ARTIC SARS-CoV-2 gene-specific primer set was synthesized by IDT. PCR amplicons were purified with Beckman Coulter AMPure XP beads and quantified by PicoGreen assay. 150ng of tiled PCR amplicons per sample was used to prepare sequencing libraries using IDT Lotus DNA Library Prep Kit. The size of the final library construct was determined on the PerkinElmer LabChip GX system and quantification was performed by qPCR with Kapa Library Quantification Kit (Roche Diagnostics. Sequencing was performed on Illumina NovaSeq 6000 S4 flow cells using 151 bp paired-end sequencing reads according to Illumina protocols.

### SARS-CoV-2 whole genome sequence analysis

Consensus SARS-CoV-2 genomes were generated from read data using a reference-based assembly pipeline (https://github.com/broadinstitute/viral-pipelines, Lemieux et al. 2020) with NC_045512.2 as the reference (Wu et al., 2020). We then filtered sequences based on quality summaries implemented in Nextclade using default parameters for SARS-CoV-2 (Aksamentov et al., 2021, https://clades.nextstrain.org). Sequences were given a “bad” missingness quality designation if they had over 3,000 sites missing. After removing sequences with a “bad” missingness score, no sequences with “bad” scores on any other quality metric remained. Of the 1,151 patients with isolates submitted to viral sequencing, 988 remained after this quality filtering. Of the 988 isolates, an additional 118 were excluded because they lacked clinical severity data. A single isolate was dropped because its viral clade designation (20A) conflicted with its position in principal component space (20I, Alpha V1), representing possible sequencing errors or a patient coinfected with multiple SARS-CoV-2 strains. The final patient sample used in subsequence analysis consisted of 869 COVID-19 patients, 106 (12.2%) of whom had severe COVID-19.

We determined the functional consequences of mutations, relative to the reference genome, by assigning them to protein-coding sequences in the viral genome and asking whether they resulted in an amino acid change (missense) or not (synonymous). Non-overlapping protein-coding sequences in the viral genome were identified by downloading 24 amino acid sequences from the UniProt PODTD1 track, which were then compared to the reference genome (The UniProt Consortium 2021, https://www.uniprot.org/uniprot/P0DTD1). Tblastn was used to obtain coordinates within the viral genome (Gertz et al. 2006). To eliminate any ambiguity in the functional consequences of mutations, we ignored codons where more than one mutation was detected in at least a single individual. We also masked known problematic sites within the SARS-CoV-2 genome (De Maio et al. 2020, https://github.com/W-L/ProblematicSites_SARS-CoV2/). For the purposes of genetic association and phylogenetic analyses, we ignored all insertions, deletions, and premature stop codon mutations. A four nucleotide deletion which leads to a premature stop codon removing the last 20 amino acids of ORF3a was detected in 30 samples. Multiple mutations resulting in premature stop codons in ORF8 were detected, indicating that this gene is not essential for the virus (Jungreis et al. 2021). A custom script was written to perform annotations.

### Viral phylogenetic and population structure analysis

We assigned samples to phylogenetic clades using Nextstrain clade definitions as implemented in the Nextclade software (Aksamentov et al. 2021, https://clades.nextstrain.org). While all samples we analyzed were from Bahrain, this tool compared our sequences to a sample representative of global viral genetic diversity. We also summarized the overall genetic structure in our sample by conducting a principal component analysis (PCA) using the prcomp function in R (R Core Team, 2022). We did not variance-scale genotypes, so principal components (PCs) should be interpreted as reflecting patterns of common genetic variation.

### Associations of host variables and viral genetics with COVID-19 severity

The following non-genetic host variables were included in the multivariable logistic regression models associating host and viral factors with disease severity: age, gender, sampling time, and vaccination status (all but one patient received the Sinopharm vaccine) (Al Kaabi et al., 2021). We also included nationality, and geographical region, reasoning that these two variables might capture socioeconomic and ethnic differences predictive of access to care and COVID-19 severity (Yancy 2020, Guha et al. 2020). Sampling time was included in order to capture potential temporal changes in the patient population or treatment that may have impacted the probability of developing severe COVID-19. We grouped nationalities into Bahraini, South Asian, and other, while the geographic region of patients was encoded as one of four governorates (Capital, Muharraq, Northern, and Southern). We also analyzed a set of comorbidities that included individual predictors for hypertension, diabetes, and smoking based on previous literature implicating these characteristics with increased disease severity. We pooled additional comorbidities that were individually rare into an ‘other comorbidity’ variable comprising immunocompromising disease, renal disease, chronic lung disease, cardiovascular disease, sickle cell disease or trait, and cancer. Throughout the text we include vaccination status when referring to comorbidities.

Sampling date, age, and gender were missing for a small number of patients with good quality sequencing data. In order to retain these individuals in the data set, sampling date was imputed as the median date within the Nextstrain-assigned cluster, age was imputed as the median overall age, and gender was imputed by coding as 0/1 and setting missing individuals to the mean value.

In all analyses presented here, we used logistic regression models to evaluate the association between genetic and non-genetic variables and the severity of COVID-19 cases. Wald tests were then used to assess statistical significance at the nominal threshold of 0.05. Non-genetic variables were analyzed first in the absence of any viral genetic information, such as Nextstrain clade assignments and PCs. Age, gender, sampling date, and vaccination status were considered baseline predictors and included in every regression model presented here. All demographic variables and comorbidities were tested individually, as well as jointly in a full model containing all non-genetic factors. The observed effects of comorbidities were qualitatively compared to findings from other populations to assess whether directions were concordant with published results. All non-genetic variables were also included as covariates in viral genetic models even if associations with severity were not statistically significant.

To analyze the association between viral population structure and COVID-19 severity, we used SARS-CoV-2 clades/’variants of concern’ as designated by Nextstrain, as well as the first seven principal components (PCs) of the genetic relatedness matrix. Because some clades appeared at low counts in our sample, for association analyses we grouped low-frequency clades together. Clades 20C, 20D, 20E (Theta), 20G, 20H (Beta, V2), 21D (Eta), and 20E (EU1) were grouped together into an ‘Other’ category. Population structure variables were also included as covariates in subsequent individual-mutation and gene-based tests. The effects of individual, common mutations were analyzed in the same manner. We tested the top 50 most frequent mutations in the sample to assess whether their presence or absence was associated with COVID-19 severity. Statistical significance was evaluated using a Bonferroni correction and family-wise error rate of 0.05.

We examined the relationship between vaccination and clades in our patient panel by grouping patients by vaccination status and clade for the months of January, February, and March 2021 (when vaccination became common). We balanced the data for each month by randomly dropping patients so that equal numbers of vaccinated and non-vaccinated individuals remained within each month. After pooling the data across the three months, we conducted a Fisher’s Exact Test between vaccination status and a binary variable indicating Alpha versus non-Alpha clade status.

The connection between individual viral genes with COVID-19 severity was investigated using a burden approach. For a given gene, each patient was assigned a viral burden score based on the number of rare mutations detected in that gene. We then tested whether patients with higher burden scores in a particular viral gene are more or less likely to have severe disease. This approach is capable of detecting severity-influencing viral genes when mutations in those genes tend to influence risk in the same direction. Given the existing evidence for negative selection in SARS-CoV-2 (Morales et al. 2021), we hypothesized that at least in certain genes, mutations which decrease viral fitness could improve patient outcomes. To enrich for mutations under negative selection, and also to avoid some of the aforementioned confounding issues with common mutations, we calculated burden scores using a frequency cutoff. Mutations under this cutoff were included in the score, while those over were ignored. We varied the frequency cutoff between 1 and 100 to assess sensitivity.

The association between burden scores and COVID-19 was tested using logistic regression, including all demographic and health information, as well as the top seven principal components (PCs) and Nextstrain clade designations, as covariates. We calculated burden scores for all missense and all synonymous mutations. In analyses of missense variation, we also included as covariates the burden score for synonymous mutations with the same frequency cutoff and for the maximum frequency cutoff (100). The purpose of including synonymous burden scores as a covariate was to correct for any overall correlation of burden with severity unrelated to function. We first tested the RNA-dependant RNA polymerase (*Pol*) and S glycoprotein (*Spike*) genes, as both are large genes with well-understood biological functions and assessed statistical significance at the nominal 0.05 level. We then calculated and tested burden scores for the remaining 12 genes. This second set contained many small genes for which we expect to have much lower power, and statistical significance was assessed using a Bonferonni-corrected threshold of 0.0024 (family-wise error rate: 0.05). To assess whether regression results were due to the composition of our sample rather than gene-specific effects, we also simulated burden scores by randomly sampling missense mutations with the same observed frequencies for each gene. Regression analyses were then performed on simulated scores, and we calculated the percentile of each gene’s effect size in the real data within this simulated distribution of frequency-matched, hypothetical genes.

We investigate the predictive value of viral genetic variables for COVID-19 severity using a leave-one-out cross-validation (LOOCV) approach. This was done by leaving out every patient from the data set one at a time, fitting a logistic model on the remaining samples, and using the fit model to predict the probability that the left-out patient developed severe COVID-19.

Hypertension, smoking status, gender, and date were left out of LOOCV regressions because they did not provide any predictive value. Regressions were fit with and without each viral genetic variable at each iteration. Predicted probabilities of severe disease were compared to true disease status by calculating false positive and false negative rates at different probability thresholds (receiver operating characteristic (ROC) analysis) and computing the area under this curve (AUC). Confidence intervals were obtained by bootstrapping patients and performing the LOOCV procedure on bootstrapped samples. Prediction analyses were performed for the first principal component and for missense burden scores in the Pol and Spike genes.

### Mutation frequency across the SARS-CoV2 genome

To explore the connection between negative selection and gene-based severity associations, we used two metrics to measure the tendency of genes to harbor lower or higher frequency mutations compared to the overall viral genome. The first metric is the average number of missense mutations observed in that gene across patients. The second is a rank percentile sum statistic (RPSS) designed to be less sensitive to the presence of outlier high-frequency, positively selected mutations. RPSS was calculated by first ranking the mutations in a gene from highest to lowest count. The percentile of each count within the set of all observed mutations was then computed, conditional on the count of the mutation with the next highest rank. The sum of these percentiles was the RPSS. For instance, for a gene with 10 mutations where the highest frequency mutation has count 100 and the next highest frequency mutation has count 50, the first percentile score would be the probability that the highest frequency was 100 or greater in a sample of 10 mutations from across the whole genome. The second percentile score would be the probability that the highest frequency mutation in a sample of size 9 was 50 or greater after all mutations with count greater than 100 are removed, as well as the single 100-count mutation in the previous spot. When the count reaches one, we stop adding percentiles. This metric therefore reflects the tendency of mutation counts within a gene to decrease by rank, compared to that expected by chance. As a rank-based statistic, RPSS should be less sensitive to the complex phylogenetic structure which could result from sampling SARS-CoV-2 patients. The RPSS strategy is similar to generalized summary statistics for the site frequency spectrum (Achaz 2009) with equal weights given to ranks rather than frequencies.

Both selection scores were compared to random samples with the same number of mutations as observed in that gene. A gene’s percentile within these samples reflects how often we would see such a bias towards low-frequencies if mutations were assigned randomly to genes within the SARS-CoV-2 genome. Because this procedure conditions on the observed number of mutations in a gene, it should reflect whether observed mutations are at lower frequencies than expected by chance, rather than overall levels of variation that are sensitive to differences in mutation rate.

## Data availability

All code for processing patient information and viral genetic data for running analyses is available at https://github.com/emkoch/CV19BH.

## Acknowledgements

We would like to thank Luca Freschi for his assistance in the early stage of this project, as well as members of the Sunyaev lab for their constructive feedback. This study was funded by an award from the Bahrain Ministry of Health to AH (Fund #102075). AH was supported by the US National Heart, Lung, and Blood Institute (NHLBI K8HL150284), and the American Heart Association. EK and SH were supported by the US National Institutes of Health grant R35GM127131 to SH.

## Supporting information

**Figure S1.**
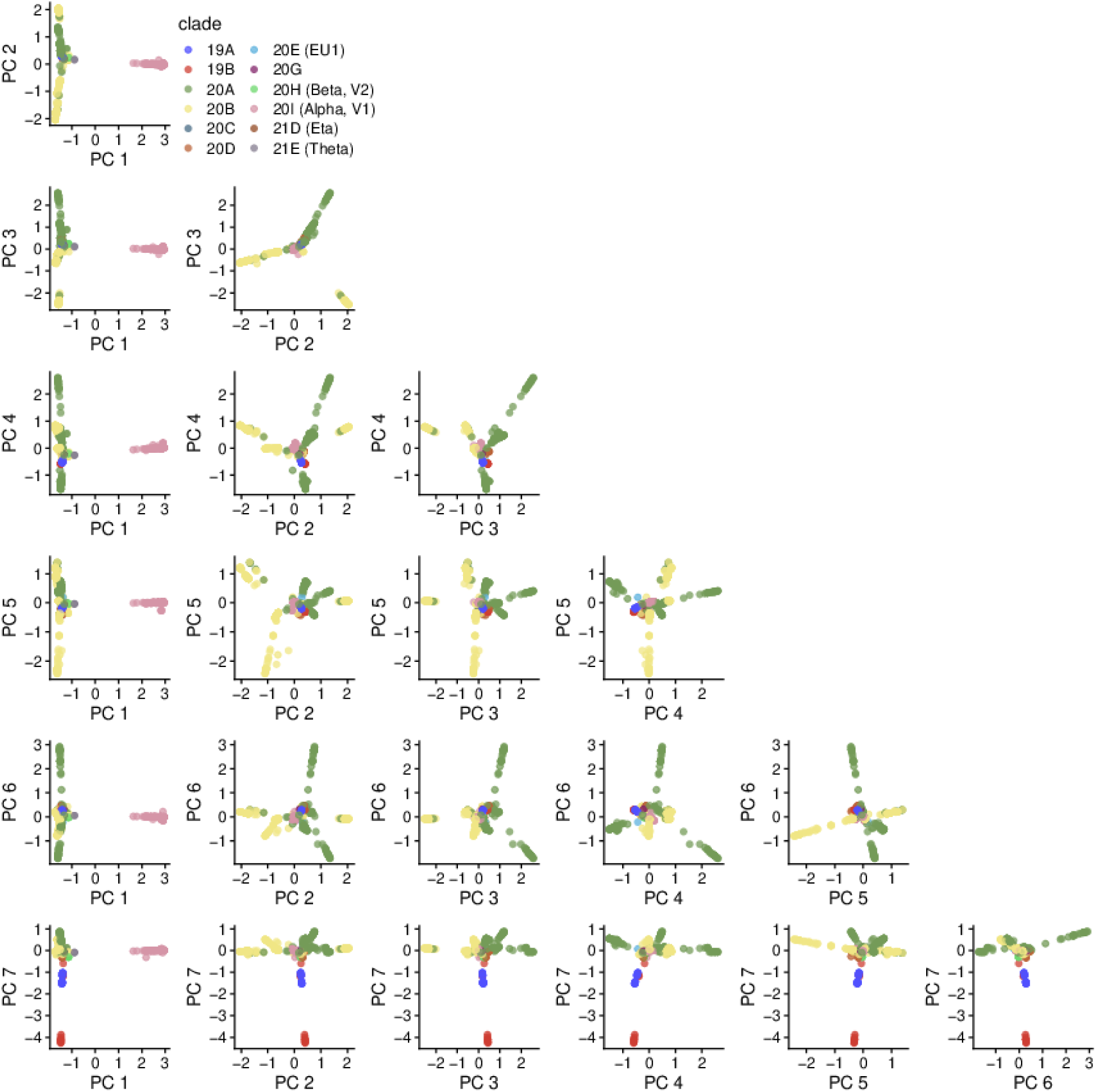
The clustering of viral genomes along the first seven PCs. PC 1 separates out clade 20I (Alpha, V1), PCs 2-6 largely capture variation within clades 20A and 20B, and PC 7 separates out 19A and 19B from the other clades.

**Figure S2.**
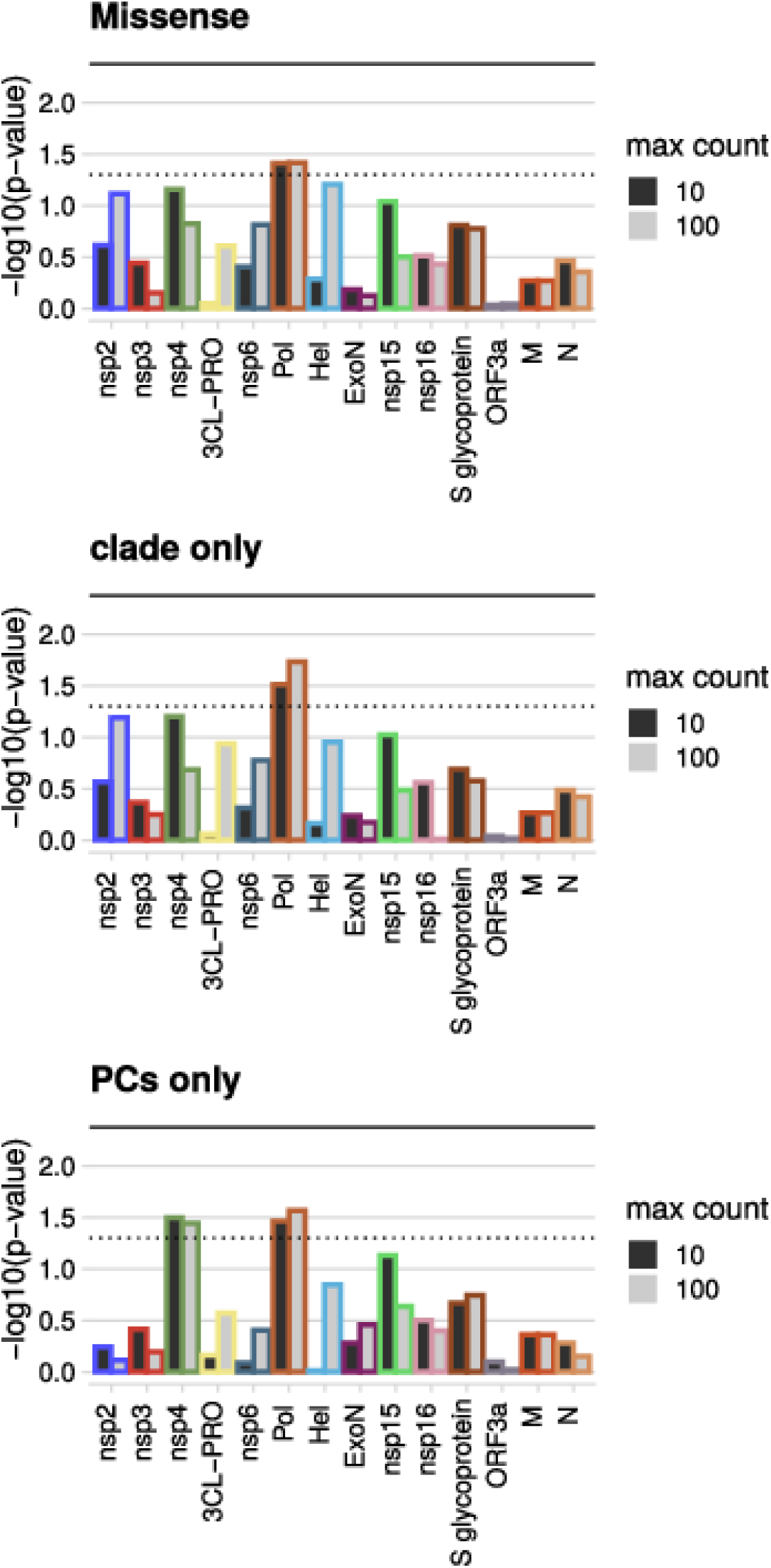
The robustness of gene-based association results to different ways of controlling for common mutations. P-values from logistic regression analyses of COVID-19 severity on burden scores in SARS-CoV-2 genes. Dotted line shows the nominal significance threshold of 0.05, and the bold line shows the Bonferonni threshold of 0.004. Burden scores of missense variants are compared to scores for synonymous variants. The top row shows results from regressions using both clades and PCs as covariantes. The next two rows show results where only either clades or PCs are included.

**Figure S3.**
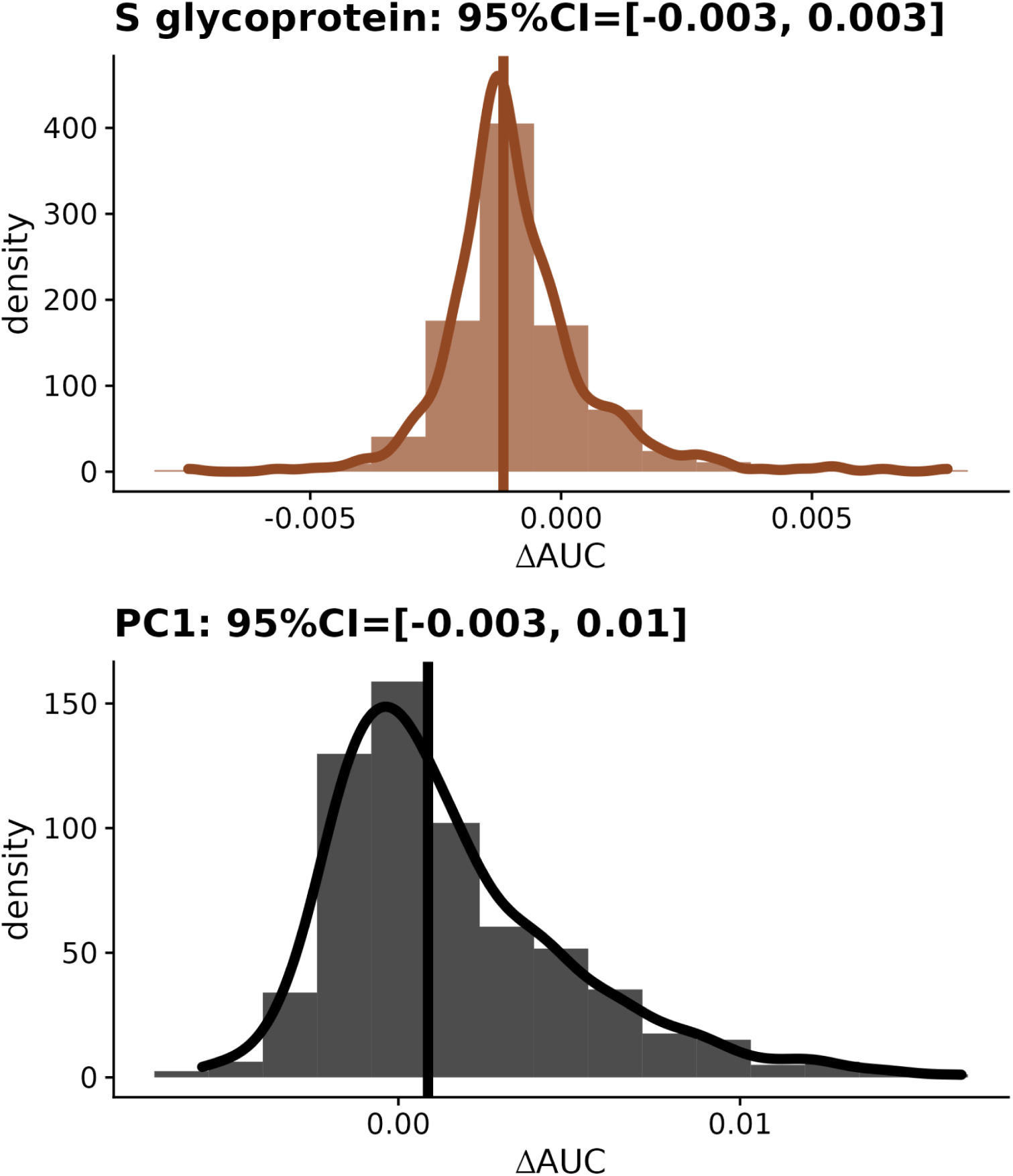
Changes in AUC resulting from adding Spike max-10 burden score and PC1 to predictions of COVID-19 severity from logistic regressions. Vertical lines show the 𝚫AUC=0.008 95%CI=[0.001, 0.017]), no prediction improvement adding AUC calculated by applying LOOCV to the full patient data. Distributions show 𝚫AUC=0.008 95%CI=[0.001, 0.017]), no prediction improvement adding AUC values calculated by applying LOOCV to bootstrap samples from the patient data. 95% confidence intervals are calculated from bootstrap samples.

